# Asymptomatic COVID-19 Have Longer Treatment Cycle Than Moderate Type of Confirmed Patients

**DOI:** 10.1101/2020.05.16.20103796

**Authors:** Wei Zhang, Qinying Long, Yanbiao Huang, Changju Chen, Jinhua Wu, Yang Hong, Hourong Zhou, Weidong Wu

## Abstract

**Objectives:** A kind of pneumonia caused by unknown causes that occurred in Wuhan, Hubei, China in December 2019, was reported as a result of novel coronavirus infection on January 7, 2020, and then WHO named it COVID-19. To compare the difference of epidemiology and clinical characteristics between asymptomatic COVID-19 infections and moderate type of confirmed cases.

**Methods:** Retrospective, single-center cohort study of COVID-19 involving 52 infections of both 26 asymptomatic and 26 moderate type of confirmed cases in the recovery stage at Guizhou Provincial Staff Hospital in Guiyang, China, from January 29, to March 31, 2020; final date of follow-up was April 22. This study was registered in Chinese Clinical Trial Registry Center. Documented the asymptomatic COVID-19 infections and moderate type of confirmed cases. Epidemiological, demographic, clinical, laboratory, radiological, and treatment data were collected and analyzed. Epidemiological and clinical characteristics of asymptomatic COVID-19 infections and moderate type of confirmed cases were compared.

**Results:** The median treatment cycle of asymptomatic COVID-19 infections was 16 days (interquartile range, 11-20 days) and longer than 13 days (interquartile range, 10-15 days) of moderate type of confirmed cases (*p*=0.049). The median incubation period of asymptomatic COVID-19 infections was 10 days (interquartile range, 0-21 days), while the control group was 7 days (interquartile range, 1-15 days) (*p*=0.27). On the initial chest computerized tomography (CT) check, 18 (69.2%, 18/26) asymptomatic COVID-19 infections were no imaging changes, which was of no significance compared with 12 (46.2%, 12/26) patients with moderate type of confirmed patients (*p*=0.092).

**Conclusions:** In this single-center study, we found that asymptomatic COVID-19 infections have longer treatment cycle than those moderate type of confirmed cases.

**Key Points:** In this single-center case series involving 52 infections with asymptomatic and moderate type of COVID-19 cases, asymptomatic COVID-19 infections have longer treatment cycle than those moderate type of confirmed patients.

---

A kind of pneumonia caused by unknown causes that occurred in Wuhan, Hubei, China in December 2019, was reported as a result of novel coronavirus infection on January 7, 2020^1,2^, and then World Health Organization named it coronavirus disease 2019 (COVID-19) on February 11^3^. As of May 1, 2020, 0:00-24:00, there were 989 cases of asymptomatic COVID-19 in China under medical observation.^4^ *The New Coronavirus Pneumonia Prevention and Control Program in China (The Fifth Edition)* clearly stated that asymptomatic infections may be a source of infection.^5^ Experts pointed out that asymptomatic COVID-19 infectors bring challenges to epidemic prevention and control due to their lack of clinical manifestations and the continuous reproduction of SARS-CoV-2 in vivo and their release to the outside world, which made them an important source of infection^6^. However, it is unclear what the difference of epidemiological and clinical characteristics between asymptomatic COVID-19 infections and moderate type of confirmed patients are. We aim to analyzed and compare the difference of epidemiological and clinical characteristics between asymptomatic COVID-19 infections and moderate type of confirmed cases via the COVID-19 clinical database of Guizhou province, China.

## Methods

### Design

This is a retrospective, single-center cohort study of COVID-19 involving 52 infections of both 26 asymptomatic and 26 moderate type of confirmed cases in the recovery stage at Guizhou Provincial Staff Hospital in Guiyang, China, from January 29, to March 31, 2020; final date of follow-up was April 22. This study was registered in Chinese Clinical Trial Registry Center.

### Setting

A tertial hospital of COVID-19 medical center in Guizhou Province.

### Patients

Inclusion criteria: 26 asymptomatic COVID-19 infections and 26 moderate type of confirmed patients. Exclusion criteria: Non-COVID-19 infections. All data were obtained from the records of a hospital information system (HIS) in Guizhou Provincial Staff Hospital. Despite the retrospective study design involving electronic health records and no additional interventions, written informed consent was required from those patients or their relatives. This study was approved by the Ethics Committee of Affiliated Hospital of Zunyi Medical University. This study was registered at the Chinese Clinical Trial Register (CCTR number: ChiCTR2000032770, registered 10 May 2020). URL: http://www.chictr.org.cn/edit.aspx?pid=53228&htm=4.

### Exposures

Documented the asymptomatic COVID-19 infections and moderate type of confirmed patients.

### Case Definition

According to *the sixth edition of New Coronavirus Pneumonia Prevention and Control Program in China*^*7*^, asymptomatic COVID-19 infections refer as those susceptible population who have been infected with SARS-CoV-2 without clinical symptoms (such as fever, cough, sore throat, etc.), but the RT-PCR of swab throat test appeared a positive results. The confirmed COVID-19 case is defined as those susceptible population were infected by SARS-CoV-2 and appeared some specific clinical manifestations, such as fever, dry cough, sore throat, anorexia, diarrhea, etc., as well as the positive changes of chest computed tomography (CT) check and RT-PCR of swab throat test. Moreover, according to the severity of the disease, the confirmed COVID-19 cases are divided into four categories: mild type, moderate type, severe type, and critical type.

#### Main outcomes and measures

Epidemiological, demographic, clinical, laboratory, radiological, and treatment data were collected and analyzed. Epidemiological and clinical characteristics of confirmed COVID-19 infections and asymptomatic infections were compared.

### Statistical Analysis

Categorical variables were described as frequency rates and percentages, and continuous variables were described using mean, median, and interquartile range(IQR) values. Means for Continuous variables were compared using independent group *t* tests when the data were normally distributed; otherwise, the Mann-Whitney test was used. Data (non-normal distribution) from repeated measures were compared using the generalized linear mixed model. Proportions for categorical variables were compared using the χ2 test, although the Fisher exact test was used when the data were limited. All statistical analyses were performed using SPSS version 13.0 software (SPSS Inc). For unadjusted comparisons, a 2-sided α of less than .05 was considered statistically significant. The analyses have not been adjusted for multiple comparisons and, given the potential for type I error, the findings should be interpreted as exploratory and descriptive.

## Results

The CONSORT Flow Diagram of this study unfolded in Figure 1. One hundred and forty six medical record number were selected by researcher. After we deleted the duplication of those number, there were 135 cases with COVID-19 infections. Twenty-six asymptomatic COVID-19 infections was regarded as the observation cohort, and another 26-moderate type of confirmed cases from those leaving 109 patients with confirmed COVID-19 infections formed a controlled cohort. We compared the epidemiological and clinical features between the two cohorts.

**Figure 1.**
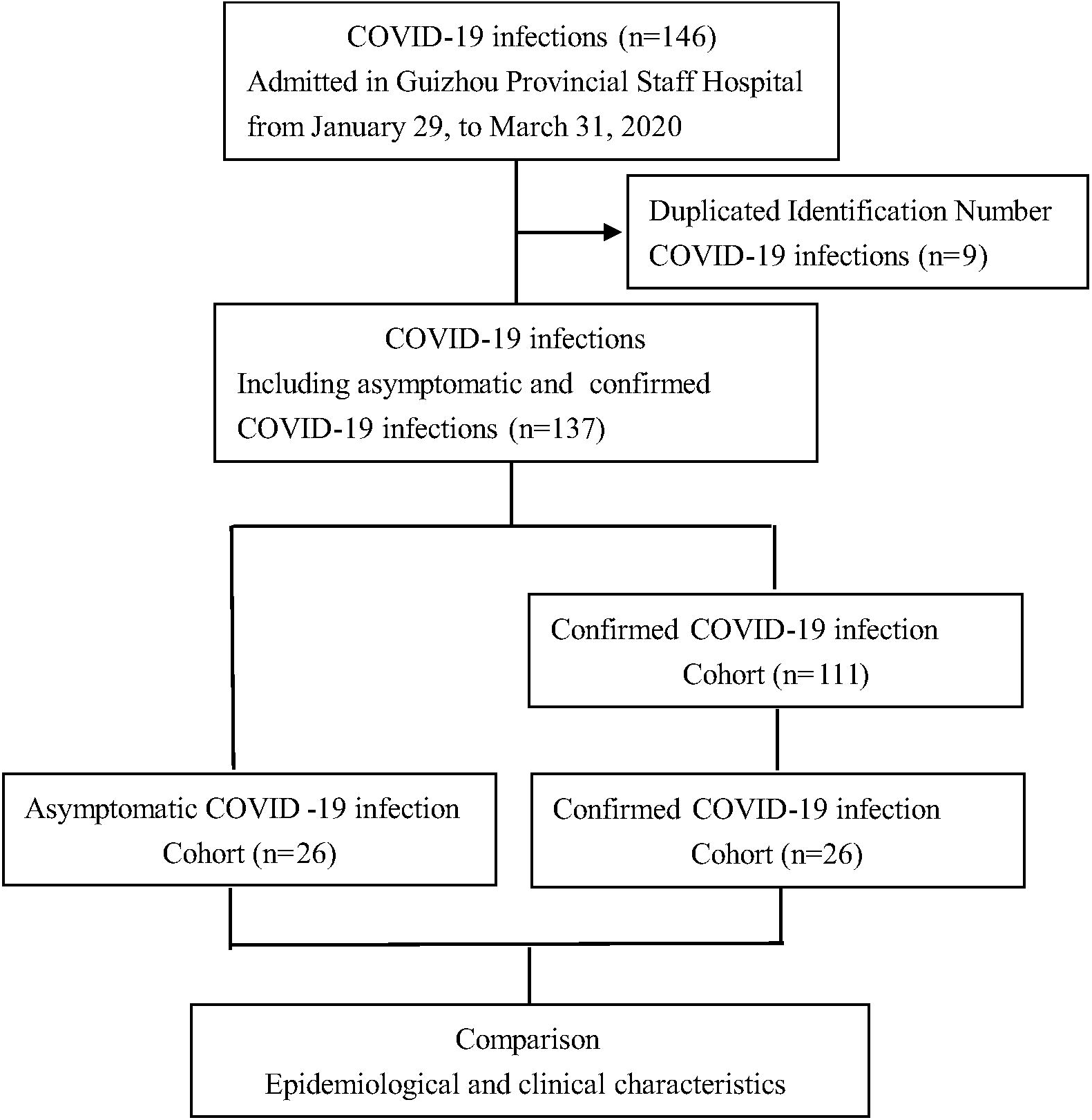
CONSORT Flow Diagram of This Study.

### Baseline characteristics

Twenty-six asymptomatic COVID-19 infections were regarded as an observation cohort. In this cohort, there were 14 male patients and account for 53.8% (14/26), and the median age was 24 years (interquartile range, 12-36 years). In racial composition, 25 COVID-19 infections belong to Han nationality (96.2%), and only one case is minority. Twenty-two (84.6%, 22/26) cases lived in Guizhou province, and 4 (15.4%, 4/26) lived in Wuhan and other cities in Hubei. Eleven cases (42.3%, 11/26) of COVID-19 infections had a history of living in situ. There were 19 cases (73.1%, 19/26) of COVID-19 infections with a familial clustered onset with median incubation of 10 days (interquartile range, 0-21 days). Median treatment cycle of 16 days (interquartile range, 11-20 days). Eighteen COVID-19 infections (69.2%, 18/26) had no clinical changes in the first check of chest CT image, 8 cases (36.4%, 8/26) showed an increase in lymphocyte ratio, 6 cases (27.3%, 6/26) had a decrease in neutrophil ratio, and a decrease in leukocyte count.

Age and gender were regarded as the matching factors, the same number of moderate type of confirmed COVID-19 infections were selected as a controlled cohort. The main onset symptoms of those COVID-19 infections were fever, and had 7 cases (26.9%, 7/26), cough 6 cases (23.1%, 6/26), expectoration 4 cases (15.4%, 4/26). In this cohort, there were 12 male infections and accounted for 46.2% (12/26). The median age was 24 years (interquartile range, 12-35 years), and there was no significant difference (*p*=0.92) for age difference analysis between the cohort of asymptomatic and moderate type of confirmed COVID-19 infections. Of the 25 cases (96.2%, 25/26) of Han nationality, 20 cases (76.9%, 20/26) of infections lived in Guizhou province, 14 cases (53.8%, 14/26) had no history of epidemic in situ, 18 cases (69.2%, 18/26) were familial clustered onset, median incubation period was 7 days (interquartile range, 1-15 days), median treatment period was 13 days (interquartile range, 10-15 days). Twelve COVID-19 infections (46.2%, 12/26) had no clinical changes in the first check of chest CT, and the abnormal laboratory examination indexes of those COVID-19 patients were rare, mainly focused on platelet count and erythrocyte sedimentation rate, and we also found that 4 cases (17.4%, 4/26) had an increased lymphocyte ratio.

### Analysis of Variances

The age, incubation period and treatment cycle of the metrological data of 52 infections were tested normally, which was skewed distribution. The differences in epidemiological and clinical characteristics between moderate type of confirmed cases and asymptomatic infections were compared (Table 1). The statistical results showed that the treatment cycle of asymptomatic cohort was significantly longer than moderate type of confirmed cohort (*p* =0.049).

### Treatment cycle

The average treatment cycle of infections with moderate type of confirmed COVID-19 patients was reported to be 14 days and within^8^. In this research, the median treatment cycle of asymptomatic COVID-19 infections was 16 days (interquartile range, 11-20 days), while the median treatment cycle of controlled group was 13 days (interquartile range, 10-15 days). The treatment cycle of asymptomatic COVID-19 infections was longer than that of confirmed moderate type of infections, and the difference between the two groups was significant (*p*=0.049).

### Incubation period

Studies have shown that the average incubation from COVID-19 possible exposure time to the first onset of the disease was 5 days^9^. In this study, the mean incubation of asymptomatic COVID-19 infections was 12 days, median incubation was 10 days (interquartile range, 0-21 days), and median incubation of controlled group was 9 days, median incubation 7 days (interquartile range, 1-15 days). In this study, the incubation period of asymptomatic infections was longer than that of previous studies, and the incubation period of COVID-19 asymptomatic infections was also longer than that of moderate type of confirmed infections, but there was no significant difference between the two groups (*p*=0.273).

### Initial check of chest CT

Chest CT check has a high diagnostic value in the diagnosis of COVID-19 pneumonia^10-13^. Whereas in this study, the first chest CT check of 18 (69.2%, 18/26) asymptomatic COVID-19 infections had no clinical changes, while the moderate type of confirmed infections had 12 cases (46.2%, 12/26). Asymptomatic COVID-19 infections had fewer imaging changes after infection with SARS-CoV-2 than confirmed infections. However, there was no significant difference between the two groups (*p*=0.092).

### Laboratory data

In view of laboratory test of asymptomatic COVID-19 infections, 8 cases (36.4%, 8/22) had an increased lymphocyte ratio, 6 cases (27.3%, 6/22) had leukopenia, 4 (18.2%, 4/22) had a decreased lymphocyte ratio and increased platelets, 3 cases (15.8%, 3/19) occurred in erythrocyte sedimentation rate (ESR) more than 20 mm per hour; while in the controlled group, 5 cases (21.7%, 5/23) had increased platelet count and 4 cases (17.4%, 4/23) had increased lymphocyte ratio. High lymphocyte ratio, neutropenia and leukopenia all showed clinical laboratory characteristics of COVID-19 infections. There was no significant difference in laboratory variables between the two groups (*p*>0.05).

## Discussion

In the susceptible population of SARS-CoV-2, the elderly and those who have co-morbidities including hypertension, diabetes, cardiovascular and cerebrovascular diseases, etc., were more susceptible and more likely to develop into severe or critical cases, even had higher mortality^1,14-16^. Early studies on novel coronavirus pneumonia (NCP) found that male patients were more susceptible than women^17^. Therefore, when we began to design this study, we had fully considered the influence of age and gender on the disease, and formed the controlled cohort. Factually, there was no significant difference in age and gender between the observation group and the control group (*p*=0.919 & 0.579), which proved that the baseline of the two groups was consistent.

### Treatment cycle of COVID-19 infections

In the study, we found that the treatment cycle of asymptomatic COVID-19 infections was significantly longer than moderate type of confirmed cases (*p*=0.049). The treatment cycle of asymptomatic COVID-19 infections was 16 days (interquartile range, 11-20 days), while that of control group was 13 days (interquartile range, 10-15 days). The reason that asymptomatic infections have longer treatment period than moderate type of confirmed cases are as follows: Firstly, the prolonged incubation period of asymptomatic infections was more likely cause SARS-CoV-2 to survive in the body for a long time. Secondly, due to lack of obvious clinical symptoms, it is not enough to judge the remission only by both RT-PCR of throat swab and imaging check of chest CT. Thirdly, our understanding and reorganization with the infectivity of asymptomatic COVID-19 infections is limited.

### Incubation of COVID-19 infections

In this research, the median incubation of asymptomatic COVID-19 infections was 10 days (interquartile range, 0-21 days). In Chinese coronavirus profiling, the median incubation of confirmed COVID-19 infections was 4 days (interquartile range, 2-7 days)^16^, while that of our study was 7 days (interquartile range, 1 to 15 days). The incubation period of both asymptomatic and moderate type of COVID-19 infections in the study were longer than that of the previous study. The reasons are unveiled as follows: Firstly, since Guizhou Province was a non-epidemic area, the initiation time of the prevention and control on the epidemic of COVID-19 was later than that of surrounding provinces. Secondly, most of the early COVID-19 infections did not report in time after exposure, resulting in a longer incubation period than other epidemic areas. Therefore, moving forward the time window of COVID-19 detection is an important measure to cut off the transmission path. Simultaneously, it is an important task to track and check those who confirmed and asymptomatic COVID-19 infections, as well as close contacts.

### Imaging check of chest CT

Imaging check of chest CT has a high diagnostic value in the diagnosis of COVID-19 pneumonia. However, there are also a few COVID-19 infections with positive RT-PCR of swab throat or clinical manifestations, but there is no abnormal imaging changes on the initial chest CT check^18^. In this study, the CT imaging feature of 18 asymptomatic COVID-19 infections (69.2%, 18/26) had no changes on the first check, and the number of those infections were more than moderate type of confirmed cases. This kind of imaging feature of asymptomatic COVID-19 infections may be related to the individual’s immune status, co-morbidities, and ages. Once they owned the resistance to SARS-CoV-2, they may become the asymptomatic COVID-19 infections, which led to their initial image feature not be easy to detect the positive signs of chest CT check. Just as the imaging data analysis of asymptomatic infections in a hospital in Nanjing, among 24 asymptomatic infections included in the study, 7 (29.2%, 7/24) cases had no imaging changes for the first time^19^. Certainly, we do not deny the diagnostic value of chest CT check, particularly in asymptomatic infections. After all, it is conducive to early detection, early isolation, and early treatment of COVID-19 infections.

### Laboratory detection

#### RT-PCR of SARS-CoV-2

RT-PCR test of SARS-CoV-2 has almost always been the *gold standard* for the diagnosis and cure evaluation of COVID-19 infections. Factually, we also regarded this test as an evaluation criterion of diagnosis and treatment efficacy of COVID-19 infections in this study. Both incubation period and treatment cycle were calculated according to this criterion.

#### Other laboratory examinations

Laboratory examination of patients with COVID-19 is mainly characterized by leukopenia or lymphocyte count reduction. However, according to the characteristics of early stage of COVID-19 infections, the results of RT-PCR screening in close contact with 738 infections with COVID-19 showed that only 2.70% (2/74) asymptomatic infections met this characteristics of the blood routine test.^20^ In this study, there was no significant difference between asymptomatic and moderate type of COVID-19 infections (*p*>0.05).

To sum up, the treatment cycle of asymptomatic COVID-19 infections is longer than those moderate type of confirmed cases (*p*=0.049), the incubation period of asymptomatic COVID-19 infections is longer than that of moderate type of confirmed cases, and most of them have no imaging changes of chest CT check, which brings great challenges to prevention and control of current COVID-19.

### Advantages and disadvantages of this study

#### The advantages of this study

We, for the first time, constructed this cohort study of 26 asymptomatic infections and 26 moderate type of COVID-19 patients using the COVID-19 database in Guizhou province, and compared the differences in epidemiological and clinical characteristics between the two cohorts. Secondly, the results of this study provided a good evidence for the prevention and control of global COVID-19.

#### Defects and deficiencies of this study

Firstly, this is a single-center and retrospective case-control study. Secondly, the sample size of this study is still small due to the limitation of the number of COVID-19 infections in Guizhou province, so it is difficult to estimate the overall characteristics of COVID-19 asymptomatic infections.

## Conclusions

In this single-center study, we found that asymptomatic COVID-19 infections have longer treatment cycle than those moderate type of confirmed cases.

## Data Availability

We stated that all the data and materials were true and available in the study.

http://www.chictr.org.cn/edit.aspx?pid=53228%26htm=4

## Declaration

### Ethics approval and consent to participate

We stated that written consent to participate was obtained from the parents or guardians of the minors who are under the age of 16, while patients aged 16-18 were obtained from themselves. This study was approved by the Biomedical Ethics Committee of Affiliated Hospital of Zunyi Medical University.

### Consent to publish

Not Applicable.

### Availability of data and materials

We stated that all the data and materials were true and available in the study.

### Patent Data

These patients have not been reported in any other submission by you or anyone else.

### Competing interests

All authors and their relatives have no competing interests involved in the article.

### Funding

This study was supported by Science and Technology Support Plan of Guizhou Province in 2019 (Qian Ke He Support [2019] 2834) and Science and Technology Plan of Guizhou Province in 2020 (Qian Ke He Fundamental [2020] 1Z061).

### Authors’ contributions

ZW had full access to all data in the present study and accepts responsibility for data management and accuracy of the data analyses. Study concept and design: ZW, and WWD. Acquisition and interpretation of data: LQY, HYB and CCJ. Drafting of the manuscript: LQY, ZW and HYB. Critical revision of the manuscript for important intellectual content: WJH, WWD and ZHR. Administrative, technical, or material support: HY, ZW and ZHR. Study supervision: HY, ZW and WWD. All authors agree to submission of the final version of this manuscript. ZW is the study guarantor.

## Acknowledgements

The authors thank Ping Lu, Min Yao, Zhengqiao Yang, Jianxia Fu, Yun Zhang, et.al. gave us selfish helps on the process of database construction of COVID-19 in this study.

## Disclosure

Neither of the authors has any conflict of interest to disclose.

## Notes

### Competing Interest Statement

The authors have declared no competing interest.

### Clinical Trial

This study was registered at the Chinese Clinical Trial Register (CCTR number: ChiCTR2000032770, registered 10 May 2020). URL: http://www.chictr.org.cn/edit.aspx?pid=53228%26htm=4.

### Clinical Protocols

http://www.chictr.org.cn/edit.aspx?pid=53228%26htm=4

### Author Declarations

This study was approved by the Ethics Committee of Affiliated Hospital of Zunyi Medical University.

